# Analytical and diagnostic performances of a high-throughput immunoassay for SARS-CoV-2 IgM and IgG

**DOI:** 10.1101/2020.11.20.20235267

**Authors:** Andrea Padoan, Chiara Cosma, Paolo Zaupa, Mario Plebani

## Abstract

**Abstract:** Reliable SARS-CoV-2 serological assays are required for diagnosing infections, for the serosurveillance of past exposures and for assessing the response to future vaccines. In this study, the analytical and clinical performances of a chemiluminescent immunoassays for SARS-CoV-2 IgM and IgG detection (Mindray CL-1200i), targeting Nucleocapsid (N) and receptor binding domain (RBD) portion of the Spike protein, were evaluated.

**Methods:** Precision and linearity were evaluated using standardized procedures. A total of 157 leftover serum samples from 81 hospitalized confirmed COVID-19 patients (38 with moderate and 43 with severe disease) and 76 SARS-CoV-2 negative subjects (44 healthcare workers, 20 individuals with rheumatic disorders, 12 pregnant women) were included in the study. In an additional series of 44 SARS-CoV-2 positive, IgM and IgG time kinetics were also evaluated in a time-period of 38 days.

**Results:** Precision was below or equal to 4% for both IgM and IgG, in all the studied levels, whilst a slightly significant deviation from linearity was observed for both assays in the range of values covering the manufacturer’s cut-off. Considering a time frame ≥ 12 days post symptom onset, sensitivity and specificity for IgM were 92.3% (95%CI:79.1%-98.4%) and 92.1% (95%CI:83.6%-97.0%). In the same time frame, sensitivity and specificity for IgG were 100% (95%CI:91.0%-100%) and 93.4% (95%CI:85.3%-97.8%). The assays agreement was 73.9% (Cohen’s kappa of 0.373). Time kinetics showed a substantial overlapping of IgM and IgG response, the latter values being elevated up to 38 days from symptoms onset.

**Conclusions:** Analytical imprecision is satisfactory as well as the linearity, particularly when taking into account the fact that both assays are claimed to be qualitative. Diagnostic sensitivity of IgG was excellent, especially considering specimens collected ≥12 days post symptom onset. Time kinetics suggest that IgM and IgG are detectable early in the course of infection, but the role of SARS-CoV-2 antibodies in clinical practice still requires further evaluations.

## 1. Introduction

Severe acute respiratory syndrome coronavirus 2 (SARS-CoV-2), the human-pathogenic beta-coronavirus causative agent of coronavirus disease 2019 (COVID-19), has spread rapidly across the world leading to an ongoing pandemic infecting millions of individual and thousands of deaths [1]. In addition to molecular testing for the etiological diagnosis of COVID-19, which is mainly based on the detection of viral RNA using reverse transcriptase polymerase chain reaction techniques (rRT-PCR) on nasopharyngeal swabs, laboratory diagnostics may measure specific IgM, IgG and IgA classes antibodies against SARS-CoV-2 antigens. There are several reasons to measure SARS-CoV-2 antibodies. First, to better understand the immune response to the virus as well as the kinetics and longevity of antibody response [2–4]. Second, COVID-19 convalescent plasma is one of the available treatment option and it is necessary to characterize the antibody response of each individual before accepting her/his for plasma donation [5]. Third, to assist in the diagnosis of patients who present later in their disease course or when the standard RNA detection methods are negative but clinical suspicion is high and other diseases have been ruled out [6,7]. Fourth, for epidemiological studies to determine seroprevalence in a population of high-risk subgroups of the population (e.g. healthcare workers) [8,9]. Finally, with increasing efforts focused on developing specific vaccines, measuring antibody responses is an essential component of determining vaccine efficacy [10].

Many serological immunoassays for SARS-CoV-2 antibodies have been developed, are available and have been granted with CE mark, emergency use authorization by the US Food and Drug Administration (FDA) or other regulatory organizations [11]. Serological assays are laboratory-based (both enzyme-linked adsorbent-ELISA- and Chemiluminescent-CLIA-immunoassays), point-of-care (POCT) tests recognizing the nucleocapsid (N), spike protein (S) and its S1/S2 subproteins or the receptor binding domain (RBD) with different analytical and clinical performances. More studies, therefore, are essential to compare and potentially open the way to harmonize the performance characteristics of available serological tests.

Aim to this paper is to evaluate the analytical and clinical performances of the Mindray CL-1200i chemiluminescent (CLIA) high-throughput assay for IgM/IgG SARS-CoV-2 antibodies recognizing the full length recombinant N protein and the receptor-binding-domain (RBD) fragment of S protein.

## 2. Materials and Methods

### 2.1 Analytical methods

The CL-series SARS-CoV-2 IgG and IgM assays are a two-step chemiluminescent immunoassays for detection of IgG and IgM SARS-CoV-2 antibodies in human serum or plasma (or EDTA or heparin), analyzed on the fully automated platform Mindray CL-1200i (Shenzhen Mindray Bio-Medical Electronics Co., Shenzen, China; distributed in Italy by Medical System, Genova, IT). This analytical system is featuring high throughput (up to 180 tests/h) and the first result is produced after 25 minutes. Samples react with paramagnetic microparticles coated with full length recombinant N protein from E. Coli and receptor-binding-domain (RBD) fragment from Eukaryotic cell expression system (IgM and IgG are targeting to RBD of the spike protein of SARS-CoV-2 o and the whole length of N protein, as declared by the manufacturer). Alkaline phosphatase-labeled anti-human IgG or IgM monoclonal antibodies are added to the reaction to form sandwich structure with microparticles captured anti-SARS-CoV-2 antibodies. At last, a substrate solution is added resulting a chemiluminescent reaction measured as relative light units (RLU) by a photomultiplier into the system. Following Manufacture’s inserts (P/N: 046-019557-00 v 1.0 for IgM and P/N: 046-019558-00 v 1.0 for IgG) cut-off values are: IgG positive◻>◻10 COI and IgM positive >1 COI, according to the manufacturer’s instructions. Manufacturers claimed that the calculated clinical sensitivities of IgM and IgG were 89,66% and 100%, respectively, while specificities of IgM and IgG were 93,32% and 94,92%, respectively.

### 2.2 Evaluation of method precision

Precision was evaluated by using three human serum pools of samples with different values. Precision estimations were obtained by means of triplicate measurements of aliquots of the same pool, performed for a total of three consecutive days, according to the CLSI EP15-A3 protocol [12]. Nested analysis of variance was used to estimate precision. The results obtained for precision were compared to that claimed by the manufacturer using the procedure recommended by EP15-A3. Repeatability and within-laboratory precision were in accordance with the repeatability and intermediate precision conditions specified in the international vocabulary of metrology (VIM, JCGM 100:2012) for precision estimation within a three-day period.

### 2.3 Linearity assessment

Linearity was assessed by using mixes of four samples pools, prepared with different IgM and IgG values, using serial dilution, as specified in the CLSI EP06 A:2003 guideline (paragraph 4.3.1) [13]. In brief, two serum pools with a measured IgM and IgG antibody values of 1.4 kAU/L and 18.28 kAU/L (high-level pools), respectively, were serially diluted with a low antibody values serum pools (0.05 kAU/L for IgM and 0.01 kAU/L IgG). All measurements were performed in triplicate.

### 2.4 Samples included in the study

A total of 157 leftover serum samples from 81 hospitalized COVID-19 patients (38 classified with moderate and 43 with severe disease according to the WHO interim guidance) and 76 SARS-CoV-2 negative subjects [44 healthcare workers (NHW), 20 patients with rheumatic disorders (RD), 12 pregnant women (Pr)] were included in the study (Table 1). All subjects underwent at least one nasopharyngeal swab test, analyzed by RT-PCR. Healthcare workers were considered negative (NHW) on the basis of at least three negative sequential molecular test results.

**Table 1:** Demographic characteristics of the 157 patients included in the study. Median and interquartile range (IQR), minimum and maximum of SARS-CoV-2 IgM and IgG levels were also reported.

| Groups | Gender<br>(F/M), % of F | Age<sup>#</sup><br>(mean±SD) | Mindray IgM (COI)<br>[median (IQR),<br>min/max] | Mindray IgG (COI)<br>[median (IQR),<br>min/max] |
| --- | --- | --- | --- | --- |
| **SARS-CoV-2 negative** |  |  |  |  |
| All SARS-CoV-2<br>negative subjects<br>(n=76) | 40/36 (37.6%) | 62.9±37.2 | 0.3 (0.2-0.4),<br>0.1/2.12 | 0.9 (0.4-3.0),<br>0.1-41.6 |
| NHW<br>(n = 44) | 19/25 (43.2%) | 45.0±15.8 | 0.3 (0.2-0.3),<br>0.1/0.4 | 0.9 (0.3-2.1),<br>0.1-21.4 |
| RD<br>(n = 20) | 9/11 (45.0%) | 32.3±3.08 | 0.4 (0.2-0.6),<br>0.1/2.1) | 0.6 (0.5-3.3),<br>0.1-41.6 |
| Pr<br>(n = 12) | 12/0 (100.0%) | - | 0.3 (0.2-0.3),<br>0.14/0.34 | 1.0 (0.6-5.8),<br>0.2-7.9 |
| **SARS-CoV-2 positive** |  |  |  |  |
| All SARS-CoV-2<br>Positive patients | 19/62 (32.2%) | 64.7±13.2 | 4.8 (2.1-7.9),<br>0.2/17.0 | 72.6 (36.5-103.0),<br>0.4-147.5 |
| Mod<br>(n = 38) | 11/27 (57.9%) | 68.0±12.6 | 4.2 (1.4-6.8),<br>0.2/13.9 | 66.2 (15.5-98.4),<br>0.4-132.7 |
| Sev<br>(n = 43) | 8/35 (42.1%) | 62.5±37.1 | 5.3 (2.6-8.9),<br>0.3/17.0 | 78.1 (50.1-106.1),<br>1.4-147.5 |

<sup>#</sup>Age data were not available for patients with rheumatic disorders; \*IQR = interquartile range.
SARS-CoV-2 Negative: NHW = negative healthcare workers; RD = subjects with rheumatic disease; Pr = Pregnant women; SARS-CoV-2 Positive: Mod = moderate disease; Sev = severe disease.

### 2.5 Evaluation of IgM and IgG time kinetics

Through the time period between March 18^th^ to March 26^th^, from hospital wards with hospitalized COVID-19 positive (confirmed by positive RT-PCR using nasopharyngeal swab samples) patients (n=46), a series of residual serum samples were anonymized, aliquoted and stored at -80°C. For the analyses, all samples, were thawed and heat inactivated (see above) in batch. Samples were then evaluated in the same analytical session. A total of 176 samples were collected from the 44 patients included in the study. The entire frame of measurements was subdivided in the following time periods from symptom onset (i.e., fever) (d=days): < 5d, 5-6d, 7-8d, 9-10d, 11-12d, 13-14d,15-16d, 17-18d, 19-20d, 21-22d, 23-24d, 25-26d, 27-28d, 29-30d, 31-34d and ≥ 38 d.

### 2.6 Statistical analyses

Repeatability and intermediate precision were estimated by using analysis of variance (ANOVA). An in-house developed R (R Foundation for Statistical Computing, Vienna, Austria) script for implementing the CLSI EP15-A3 protocol was used for ANOVA and for calculating the upper verification limit. For time kinetic evaluation, the following strategy was used: for each sample included in the study, the collection date was matched with the corresponding date of symptom onset (i.e., fever). The GraphPad Prism version 9.0 for Windows was used to evaluate kinetic data. The mean IgM and IgG results (and standard errors) were plotted against the time from symptom onset (treated as continuous values). Smoothing splines with four knots were used to estimate a possible fit for the time kinetic curve. Mean, Median, standard deviation and interquartile ranges (IQR) were used to describe quantitative data, while Fisher’s exact test was used to identify association between categorical variables. Stata v16.0 (Statacorp, Lakeway Drive, TX, USA) was used as statistical software, while the “diagt” module was used to estimate diagnostic performances.

### 2.7 Ethical statement

The study protocol (number 23307) was approved by the Ethics Committee of the University-Hospital of Padova. All the patients were informed of the study and voluntarily agreed to participate. All the patients who agreed to participate provided written consent.

## 3. Results

### 3.1 Imprecision

Precision results for IgM and IgG antibody Mindray CL-1200i are reported in Table 2. The results obtained for the lower levers were not reported in the table since standard deviations were too low to estimate the coefficient of variation (0.004 and 0.005 for IgM and IgG, respectively). The precision results obtained for the intermediate and the high concentration values are satisfactory, being lower than those reported by the manufacturer.

**Table 2:** Precision results for Mindray SARS-CoV-2 IgM and SARS-CoV-2 IgG antibody assays expressed as coefficient of variation (CV) in percentage (%), obtained by using pools of samples.

| Measurand | Level (AU/mL) | Design | Laboratory Evaluation – CV% |  | Manufacturer's claim – CV (%)<sup>#</sup> |
| --- | --- | --- | --- | --- | --- |
|  |  |  | *Repeatability* | *Within-Lab Precision* |  |
| SARS-Cov-2 IgM | 0.05 | 3x3<br>(CLSI Ep15-A3) | -<sup>*</sup> | -<sup>*</sup> | - |
|  | 1.39 |  | 2.10 | 2.68 | 2.62 |
|  | 2.32 |  | 1.21 | 1.99 | 3.18 |
| SARS-Cov-2 IgG | <0.01 | 3x3<br>(CLSI Ep15-A3) | -<sup>*</sup> | -<sup>*</sup> | - |
|  | 13.39 |  | 1.74 | 3.66 | 4.26 |
|  | 31.24 |  | 1.89 | 4.03 | 3.85 |

<sup>#</sup> obtained from the Mindray SARS-CoV-2 IgM insert, 046-01-9557-00 (1.0), and from SARS-CoV-2 IgM insert, 046-019558-00 (1.0).
<sup>\*</sup> indicate that standard deviations were too low to estimate the coefficient of variation (0.004 and 0.005 for IgM and IgG, respectively).

### 3.2 Linearity

Linearity results for IgM and IgG antibody Mindray CL-1200i are shown in Figure 1. For both IgM and IgG, all tested mixes of pools deviated from linearity, being the second order coefficient of the polynomial equation highly statistically significant (p < 0.01 for both).

**Figure 1:**
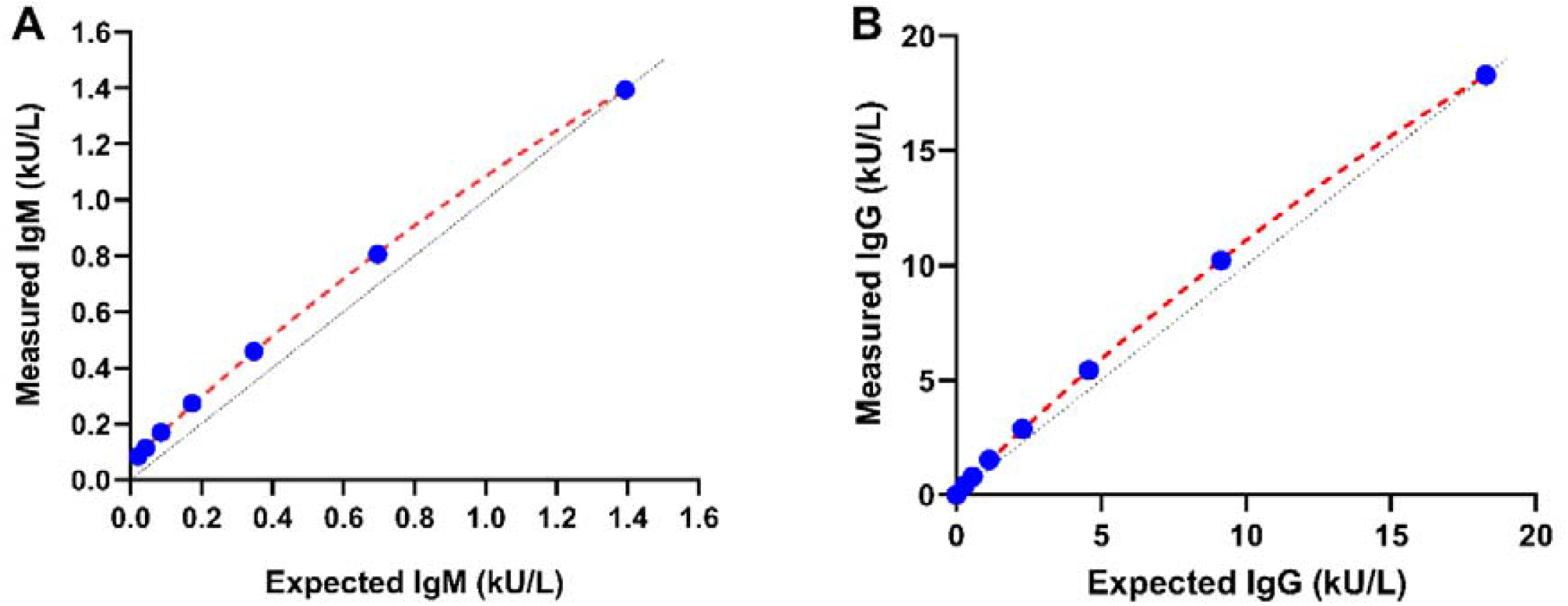
IgM and IgG antibody assay linearity results. For IgM, a high-level pool at 1.4 kU/L was serially diluted with a low-level pool at 0.01 kU/L. For IgG, a high-level pool at 18.70 kU/L was serially diluted with a low-level pool at 0.5 kU/L. Averages of triplicate measurements are shown. Second order polynomial interpolation reported a significant deviation from linearity for both IgM and IgG.

### 3.3 Demographic characteristics of individuals included in the study

Considering all samples included in the study, demographic characteristics of included individuals were evaluated. Age significantly differs between SARS-CoV-2 negative and positive subjects (Kruskall-Wallis test, χ^2^=22.1, p = 0.001). Age differences were also found among the groups of SARS-CoV-2 negative subjects (NHW vs RD, Kruskall-Wallis test, χ^2^=7.7, p = 0.005) and among SARS-CoV-2 positive subjects (Mod SARS-CoV-2 vs Sev SARS-CoV-2, Kruskall-Wallis test, χ^2^=5.4, p = 0.020). The percentages of males/females differ among groups of SARS-CoV-2 negative subjects (Fisher’s exact test, p = 0.001), but not among SARS-CoV-2 positive subjects (Mod SARS-CoV-2 vs Sev SARS-CoV-2, Fisher’s exact test, p = 0.431). The time from symptom onset and the serological determination ranged from 1 to 88 days, with a mean value of 13.1 and a standard deviation of 11 days. The time from symptoms onset did not differ by disease severity (Kruskall-Wallis test, χ^2^=2.6, p = 0.108).

### 3.4 Mindray CL-1200i IgM and IgG antibodies levels in the studied individuals

Figure 2 reports the results for SARS-CoV-2 IgM and IgG levels in all the studied groups. For IgM, significant differences in antibody levels were found between A) NHW and Mod SARS-CoV-2 (p < 0.001) or Sev SARS-CoV-2 (p < 0.001), B) between Pr and Mod SARS-CoV-2 (p < 0.001) or Sev SARS-CoV-2 (p < 0.001) and C) between RD and Mod SARS-CoV-2 (p < 0.001) or Sev SARS-CoV-2 (p < 0.001). For IgG, significant differences in antibody levels were found between A) NHW and Mod SARS-CoV-2 (p < 0.001) or Sev SARS-CoV-2 (p < 0.001), B) between Pr and Mod SARS-CoV-2 (p = 0.003) or Sev SARS-CoV-2 (p < 0.001) and C) between RD and Mod SARS-CoV-2 (p < 0.001) or Sev SARS-CoV-2 (p < 0.001). No statistical differences (p=0.999) were found between Mod SARS-CoV-2 or Sev SARS-CoV-2 patients, both for IgM and IgG. A modest correlation between Mindray CL-1200i SARS-CoV-2 IgM and gender (Spearman’s rho = 0.185, p = 0.020), as well as with age (Spearman’s rho = 0.414, p < 0.001). Similarly, SARS-CoV-2 IgG only slightly correlated with gender (Spearman’s rho = 0.264, p = 0.020), whilst they showed a moderate correlation with age (Spearman’s rho = 0.483, p < 0.001).

**Figure 2:**
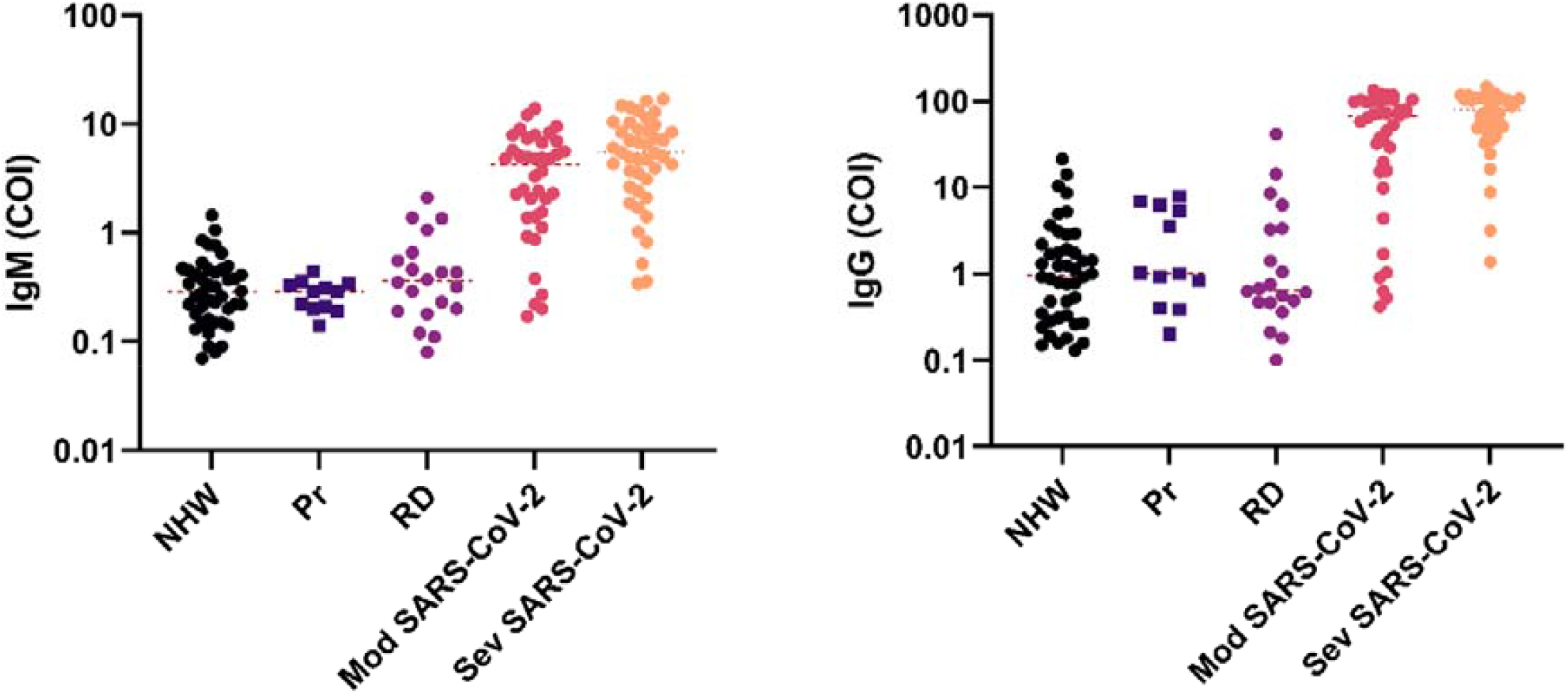
Dot-plots of Mindray CL-1200i SARS-CoV-2 IgM and IgG levels plotted with y-axes in both log_10_ scale, subdivided by studies groups (NHW = negative Healthcare Workers, Pr = Pregnant women, RD = patients with rheumatic disease, Mod SARS-CoV-2 = SARS-CoV-2 positive patients with moderate disease, Sev SARS-CoV-2 = SARS-CoV-2 positive patients with moderate disease).

### 3.5 Clinical performances of Mindray SARS-CoV-2 IgM and IgG

The numbers (and percentages) of SARS-CoV-2 IgM positive antibody tests were: NHW, n = 2/44 (4.5%), Pr, n = 0/12 (0%), RD, n = 4/20 (20.0%), Mod SARS-CoV-2, n = 31/38 (81.6%), Sev SARS-CoV-2 patients, n = 39/43 (90.7%). The numbers (and percentages) of SARS-CoV-2 IgG antibody tests were: NHW, n = 3/43 (6.8%), Pr, n = 0/12 (0%), RD, n = 2/20 (1.27%), Moderate SARS-CoV-2 patients, n = 30/38 (78.9%), Severe SARS-CoV-2 patients, n =40/43 (93.0%). Overall performances for Mindray SARS-CoV-2 IgM and IgG were estimated by the Area Under the Receiver Operating Characteristic curve (AUROC). For Mindray SARS-CoV-2 IgM and IgG, AUROC values are 0.943 (95% CI: 0.905 to 0.980) and 0.944 (95% CI: 0.908 to 0.980), respectively. Sensitivities, specificities, positive and negative likelihood ratios are reported in Table 3, estimated using overall data or considering only the period ≥ 12 days post symptoms onset. Agreement between IgM and IgG was 73.9% (percentage of concordance), with a Cohen’s kappa of 0.373, SE = 0.0731.

**Table 3:** Clinical performances SARS-CoV-2 IgM and IgG calculated considering overall data and the period ≥ 12 days post symptoms onset.

|  | **Sensitivity\***<br>**(95% CI)** |  | **Specificity\***<br>**(95%CI)** | **Positive Likelihood ratio**<br>**(95%CI)** |  | **Negative Likelihood ratio**<br>**(95%CI)** |  |
| --- | --- | --- | --- | --- | --- | --- | --- |
| | Overall | $\geq 12$ days post<br>symptom onset | Overall | Overall | $\geq 12$ days post<br>symptom onset | Overall | $\geq 12$ days<br>post symptom<br>onset |
| Mindray SARS-CoV-2 IgM | 86.4 (77.0-93.0) | 92.3 (79.1-98.4) | 92.1 (83.6-97.0) | 10.9 (5.1-23.7) | 10.8 (5.1-22.7) | 0.1 (0.11-0.32) | 0.1 (0.04-0.26) |
| Mindray SARS-CoV-2 IgG | 86.4 (77.0-93.0) | 100.0 (91.0-100.0) | 93.4 (85.3-97.8) | 13.14 (5.6-30.8) | 15.2 (6.5-35.5) | 0.15 (0.08-0.25) | 0.01 (0.01-0.21) |

\* expressed as percentages; Number of specimens evaluated: for overall data n = 157, for $\geq 12$ days post symptom onset n = 115.

### 3.6 Time kinetics of IgM and IgG

Figure 3 shows the time kinetics results at different days after fever onset, divided into time-frame categories. In detail, the plot shows average values and corresponding standard errors of IgM and IgG for each time category. Overlapping time kinetic trends are shown using spline interpolation.

**Figure 3:**
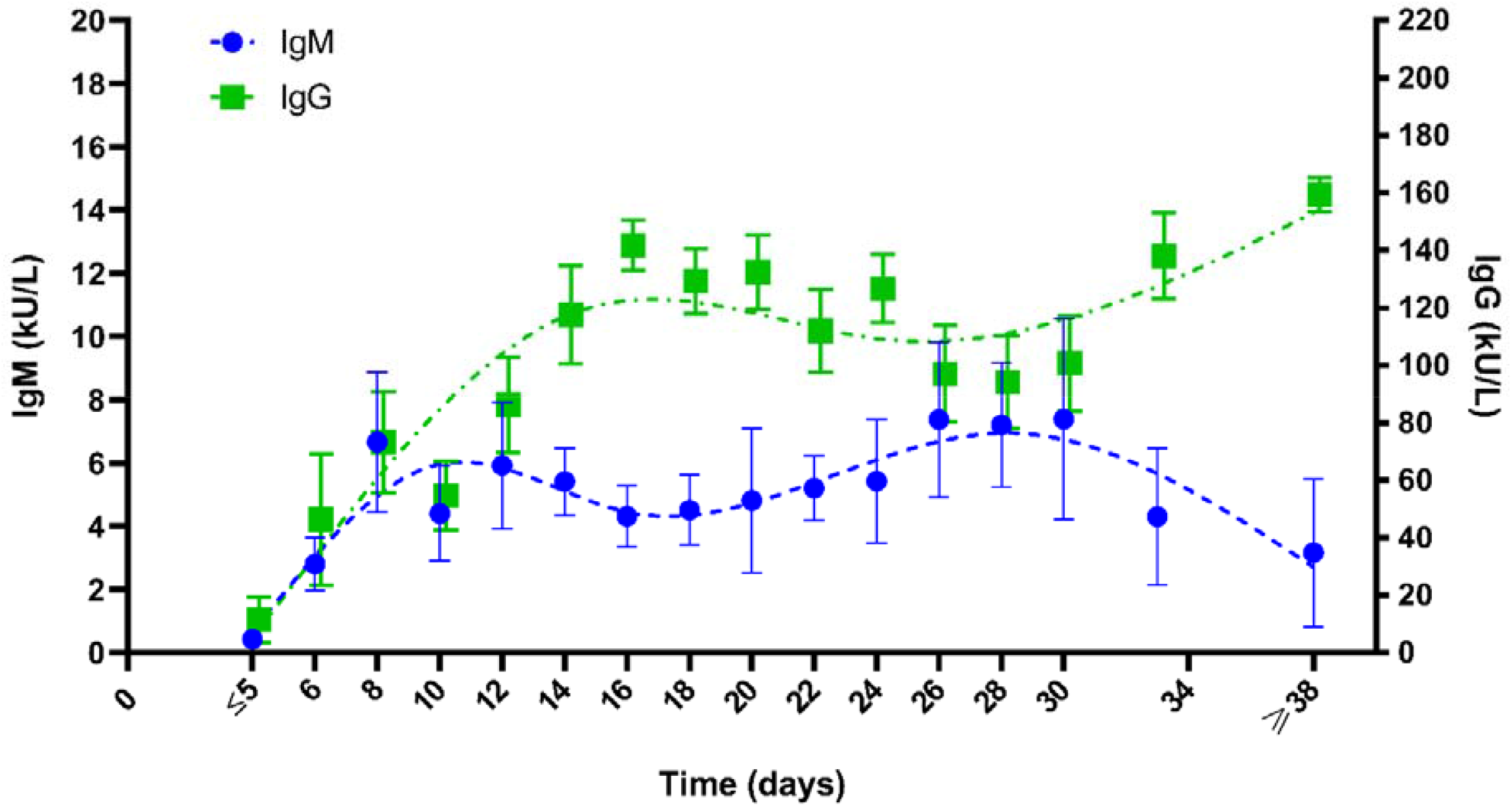
Time kinetic results of Mindray SARS-CoV-2 IgM and IgG results. In this study, a series of 44 COVID-19 positive patients (at nasopharyngeal rRT-PCR test) were included and followed-up. Mean and Standard errors are reported. Spline regressions (with 4 points knots) were used to generate the time-kinetic trends.

## 4. Discussion

Serological testing for the identification of past exposure to SARS-CoV-2 is currently of clinical and epidemiological interest with current perspective in assessing antibody response in vaccine trials. Serological testing are not well suited for the early diagnosis of COVID-19 specific antibodies being detectable after 9-12 days after symptom onset, but provides useful information for better understanding the immune-response of the individual and communities to the virus, to allow the diagnosis of patients who diagnosis of patients who present later in their disease course or experienced mild symptoms (or were asymptomatic) avoiding the standard RNA detection methods. Specific, sensitive and reliable assays are required to meet these needs. In addition, both the percentage of patients with detectable specific antibodies and the stability of the antibody response over time may also depend on the target antigen. The trimeric spike and in particular RBD, but not the nucleocapsid alone, assures a longer detectability which is well related to the persistence of neutralizing antibodies (until 148 days after symptom onset, with a range 113 to 186 days) [14].

In this study, the analytical and clinical performances of Mindray CL-1200i SARS-CoV-2 IgM and IgG assays, both targeting the N protein and RBD fragment of the S protein, were evaluated. The S protein plays an important role in specific receptor recognition and subsequent cell fusion. Hence, this protein is a major target for neutralizing antibodies. It has also been demonstrated that RBD of the S protein contains a key neutralization determinant which can induce potent neutralizing antibodies [15].

Excellent results were found for precision of both assays at clinically significant levels and the obtained results were similar to the imprecision values claimed by manufacturers. Furthermore, the precision of the lowest concentration levels was very satisfactory but estimated standard deviations were too low to derive reliable estimations. Linearity was also assessed to test the ability of Mindray CL-1200i to provide results proportional to the concentration levels. For this purpose, a series of serial dilution of high-values pools were performed. Figure 1 shows that in a range of values covering the manufacturer’s cut-off, between 1.3 COI and 0.1 COI for IgM, and between 18 COI and 0.2 COI for IgG, results were almost linear, although polynomial regressions showed a statistical significant second-order coefficient. These results were expected since both assays are claimed by the manufacturer to be qualitative, not quantitative tests.

The kinetics of IgM and IgG were also evaluated considering an additional series of serum samples from 44 positive COVID-19 patients to study the immunoglobulin response obtained after some days from infection. The results, as reported in Figure 3, show that both IgM and IgG rapidly increase after the onset of fever. Considering the cut-offs suggested by the manufacturer (i.e., 1.0 AU/mL for IgM and 1.1 AU/mL for IgG), the immunoglobulin rise could be considered significant seven days after fever onset. These findings are in agreement with those reported by us [2] and other Authors [6,16,17]. In addition, Kowitdamrong et al. showed that seroconversion is fast in patients with Mod/Sev disease than in mild disease [16]. In our time-kinetic study, in fact, all individuals were with Mod/Sev disease and further studies are requested to better understand differences in immune response in asymptomatic and paucisymptomatic patients.

Clinical performances were elevated, especially for IgG assay while IgM presents a lower diagnostic accuracy, confirming previously reported results with other immunoassays [2, 4]. Interestingly, considering samples collected ≥ 12 days after symptom onset, 100% of SARS-CoV-2 patients presented positive IgG levels, whilst two NHW and two RD individuals resulted false positive. The latter issue explain why specificity, albeit elevated (93.4%), did not achieve 100% (95%CI were 85.3% to 97.8%). The IgG sensitivity was elevated when compared with results obtained in a comparable time frame using other chemiluminescent analytical systems [4].

This study presents several limitations. First, only Mod/Sev SARS-CoV-2 individuals were included in the study, as sera from asymptomatic/paucisymptomatic subjects were not available. Second, IgM and IgG kinetics has been assessed in a limited period, while a longer monitoring should be necessary in order to estimate the entire trend of humoral immune response to COVID-19 infection, particularly the eventual antibodies decay. Third, comorbidities were not evaluated since not completely recorded.

In conclusion, Mindray CL-1200i SARS-CoV-2 IgM and IgG antibodies assays show excellent precision results and linearity, comparable to that claimed by manufacturer’s insert. Clinical performances in terms of both AUROC and sensitivities were also very elevated, especially after 12 days post symptom onset. The presence of a limited number of false positive results prevented to achieve 100% specificity, thus indicating that this point could be further improved. Finally, the overlapping time-kinetics between IgM and IgG specific antibodies, particularly in the early stages of the infection, suggests that the diagnostic value of IgM assay is still debatable.

## Data Availability

Data will be available upon request

